# A genome-wide association study in 10,000 individuals links plasma N-glycome to liver disease and anti-inflammatory proteins

**DOI:** 10.1101/2024.07.08.24309967

**Authors:** Sodbo Sharapov, Anna Timoshchuk, Olga Zaytseva, Denis Maslov, Anna Soplenkova, Elizaveta E. Elgaeva, Evgeny S. Tiys, Massimo Mangino, Clemens Wittenbecher, Lennart Karssen, Maria Timofeeva, Arina Nostaeva, Frano Vuckovic, Irena Trbojević-Akmačić, Tamara Štambuk, Sofya Feoktistova, Nadezhda A. Potapova, Viktoria Voroshilova, Frances Williams, Dragan Primorac, Jan Van Zundert, Michel Georges, Karsten Suhre, Massimo Allegri, Nishi Chaturvedi, Malcolm Dunlop, Matthias B. Schulze, Tim Spector, Yakov A. Tsepilov, Gordan Lauc, Yurii S. Aulchenko

## Abstract

More than a half of plasma proteins are N-glycosylated. Most of them are synthesized, glycosylated, and secreted to the bloodstream by liver and lymphoid tissues. While associations with N-glycosylation are implicated in the rising number of liver, cardiometabolic, and immune diseases, little is known about the genetic regulation of this process. Here, we performed the largest genome-wide association study of N-glycosylation of the blood plasma proteome in 10,000 individuals. We doubled the number of genetic loci known to be associated with blood N-glycosylation by identifying 16 novel loci and prioritizing 13 novel genes contributing to N-glycosylation. Among these were the *GCKR*, *TRIB1*, *HP, SERPINA1* and *CFH* genes. These genes are predominantly expressed in the liver and show a previously unknown genetic link between plasma protein N-glycosylation, metabolic and liver diseases, and inflammatory response. By integrating glycomics, proteomics, transcriptomics, and genomics, we provide a resource that facilitates deeper exploration of disease pathogenesis and supports the discovery of glycan-based biomarkers.

---

During maturation, more than half of human proteins are modified by the covalent linking of complex carbohydrates – glycans^1^. Glycoproteins comprise various secreted and membrane enzymes, receptors, hormones, cytokines, immunoglobulins, as well as structural and adhesion molecules^2^. Glycans affect the physical and chemical properties of proteins and their biological function^3–6^. Adequate glycosylation is required for the normal physiological action of glycoproteins, while aberrant glycosylation is increasingly implicated in human diseases^7–9^. Glycans are considered to be potential therapeutic targets^10–12^, essential part of therapeutics^13–15^, as well as biomarkers^16–18^, which makes glycobiology a promising field for future clinical applications.

N-glycosylation is the most abundant type of glycosylation^1^ and, unlike other types, is specific to a consensus asparagine-containing sequence (Asn-X-Ser/Thr, where X is any amino acid except Pro) in the protein’s primary structure. Human N-glycans are irregular branched polymers consisting of mannose, galactose, fucose, sialic acid, and *N*-acetylglucosamine (GlcNAc) residuals, whose combinations introduce a great diversity of protein glycoforms. Unlike proteins, whose primary structure is encoded in the genomic DNA sequence, the occupancy of the N-glycosylation site, and the abundance of specific N-glycan structures are not directly encoded in the human genome. Protein glycosylation depends on the interplay of multiple enzymes catalyzing glycan transfer, glycosidic linkage hydrolysis, and glycan biosynthesis. The abundance of specific protein glycoforms can be influenced by various parameters, including the activity of enzymes and availability of substrates, the accessibility of a glycosylation site, protein synthesis, and degradation^19^. Overall, protein glycosylation is a complex process controlled by genetic, epigenetic, and environmental factors^20–22^.

While the biochemical network of human N-glycan biosynthesis is well understood^23^, little is known about *in vivo* regulation of this process^24^, including tissue- and protein-specific regulation. A major part of the plasma glycoproteins consists of immunoglobulins, produced by antibody-producing B-cells, and secreted proteins produced in the liver^25^.Therefore, the N-glycosylation of blood plasma proteins serves as an indicator of liver and B-cell function. Study of plasma protein N-glycosylation potentially provides insights into the etiology and pathophysiology of liver and B-cell-mediated diseases, as well as diseases where these tissues are important players, such as cardiometabolic diseases and inflammatory conditions. Understanding the mechanisms underlying blood plasma glycosylation and its regulation at the tissue-specific level is crucial for unraveling the complex interplay between protein modifications, cellular functions, and disease processes.

In this context, genetics offers an attractive approach to studying regulation of N-glycosylation *in vivo* and sheds light on how these molecular phenotypes are linked to human disease^25,26^. Abundance of total plasma N-glycans can be quantified through various analytical methods^27^. As for other quantitative phenotypes, the genome-wide association study (GWAS) and multivariate genetic association analysis^28,29^ may be applied to N-glycans to identify genetic loci associated with abundance and, therefore, contain genes involved in the regulation of N-glycosylation. Further integration of N-glycome GWAS results with other layers of biological information (e.g., biological pathways, protein-protein interactions, transcriptomics, proteomics, and others) allows the discovery of novel candidate genes regulating this process and provides hypotheses about biological mechanisms underlying the genetic associations^30,31^. A joint analysis of GWAS results of N-glycome and disease (e.g., pleiotropy analysis^32^ and analysis of causal relationships using Mendelian randomization^33^) can shed light on how protein glycosylation is involved in pathogenesis of human disease and suggest possible glycome-based biomarkers. Previous GWAS of total plasma N-glycome^34–37^ identified 15 genetic loci and suggested the role of 19 candidate genes. These studies were supplemented with GWAS of N-glycome of immunoglobulin G (IgG)^28,38–41^ and transferrin (TF)^42^ glycoproteins, identifying an additional 19 loci and prioritizing 26 candidate genes^25^. The role of three candidate genes, encoding transcriptional factors *HNF1A*, *IKZF1,* and *RUNX3*, in the regulation of N-glycosylation was experimentally confirmed *in vitro*^34,40^. A Mendelian randomization study of IgG N-glycome found that the abundance of N-glycans with bisecting GlcNAc is a potential biomarker of systemic lupus erythematosus^43^. However, there remains a limited understanding of the role of genes-regulators of N-glycosylation in health and disease.

The first aim of this study was to identify novel glycome quantitative trait loci (glyQTLs), prioritize novel candidate genes, and reconstruct tissue-specific gene networks that regulate plasma protein glycosylation. For this, we performed the largest genome-wide association meta-analysis (GWAMA) of total plasma N-glycome using data from seven studies (N = 10,764). For replicated glyQTLs, we prioritized candidate genes using a broad spectrum of methods and explored how these genes are connected in a functional tissue-specific network that regulates protein glycosylation. The second aim of this study was to identify potential glycan biomarkers for disease. We performed a phenome-wide association study (PheWAS) in conjunction with colocalization analysis to investigate the pleiotropic effects of glyQTLs on complex diseases. Next, we correlated genetically predicted glycan levels in 450,000 UK Biobank samples with the disease’s endpoints. Finally, we conducted a bidirectional Mendelian randomization study to identify potential causal effects between glycans and disease. This strategy not only resulted in the discovery of new candidate genes but also suggested how some of these genes might regulate glycosylation enzymes and how they are linked to the aberrant glycosylation observed in disease.

## Results

### Single- and multi-trait GWASs for 138 N-glycome traits

The levels of 36 N-glycan structures (Supplementary Table 3a, 3c) linked to various plasma glycoproteins were measured by ultra-high performance liquid chromatography in seven participating cohorts from six countries. Majority of 10,764 participants (94.5%) were of European ancestry. From the 36 directly measured N-glycans, we computed 81 derived N-glycome traits such as the total level of fucosylation, galactosylation, sialylation and others, reflecting pathways of N-glycan biosynthesis (Supplementary Table 3a). We conducted GWAS for each of these 117 N-glycome traits in each of the seven participating cohorts, assuming an additive model of the genetic effect. We then performed a fixed-effect discovery meta-analysis of the subcohorts that included participants of European descent (N = 7,540) (Supplementary Table 1b). After meta-analysis, we took advantage of the correlation structure between 117 N-glycome traits and performed GWAS of 21 multivariate N-glycome traits defined based on their biochemical similarities (Supplementary Table 1b).

The size of the replication sample (N = 3,224, Supplementary Table 1b) was defined as to achieve 80% statistical power for a replication of the true association signal (Supplementary Note).

The genomic control inflation factor in the discovery GWAMA varied from 1.004 to 1.059. By contrast, an intercept of LD score regression^44^ varied from 0.996 to 1.002 (Supplementary Table 4c), confirming minimal impact of genetic stratification on the GWAS results. Hence, implementing Genomic Control correction in the analysis was unnecessary.

Our analyses identified and replicated a total of 40 loci (**Fig. 1a**, Supplementary Table 5a, Supplementary Table 5b) that were significantly associated with at least one of 117 N-glycome traits and 21 multivariate N-glycome traits. The association of 25 loci with total plasma N-glycome was shown and replicated for the first time (**Table 1**), while the association of 15 loci confirms previous findings^36,37^.

**Figure 1:**
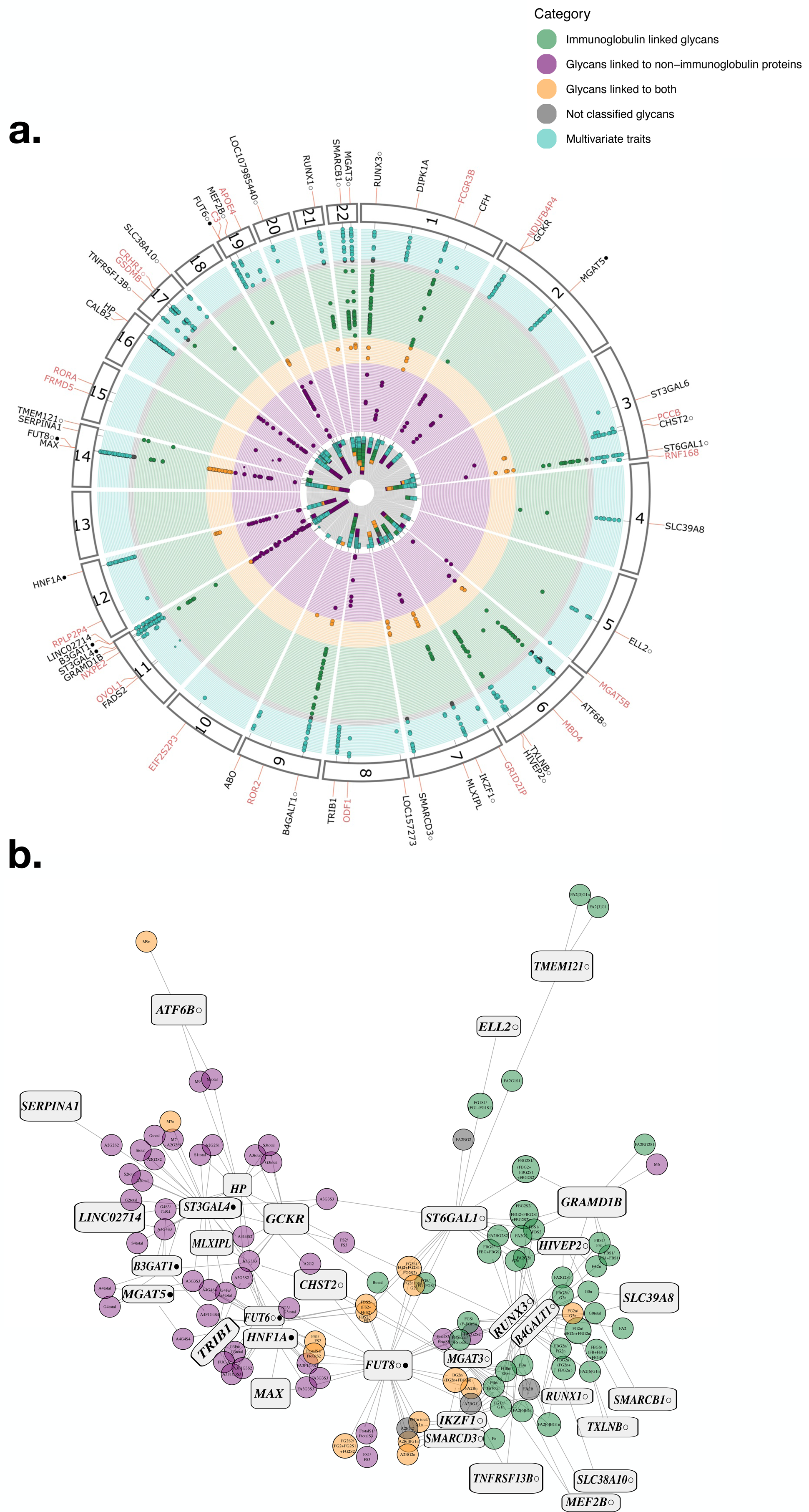
Discovered loci. (a) Associations of 59 loci with 138 glycomic traits labeled by the prioritized candidate or nearest gene names. Marked black: loci are discovered and replicated in this work; red: discovered, but not replicated in this work. In total, 117 univariate traits were analyzed (Supplementary Table 3a), but for two of them, no genome-wide significant associations were found. Univariate traits are grouped into 4 categories: glycans mostly linked to immunoglobulins (green), glycans mostly linked to non-immunoglobulin proteins (purple), glycans linked to both types of proteins (orange), not classified glycans (gray). The details of glycan classification are described in Supplementary Note. Also, the results from analysis of 21 multivariate traits (turquoise) are presented. The multivariate traits were defined based on biochemical similarities between 36 directly measured total plasma N-glycan traits (Supplementary Table 3b). (b) A network view of associations between loci and glycan traits. Rectangular nodes represent genetic loci labeled with the names of the prioritized candidate or nearest genes; circle nodes represent glycan traits. Lines represent significant genetic associations between locus and specific glycans. The colors of circle nodes are consistent with those in (a). Symbol ○ next to the candidate gene indicates that the locus was previously discovered in immunoglobulin G N-glycome GWASs; Symbol ● — the locus was previously discovered in transferrin N-glycome GWAS, as reviewed in ^25^.

**Table 1:** Twenty-five novel loci associated with total plasma N-glycosylation discovered and replicated in this study. Full results of discovery and replication are provided in Supplementary Table 5a, 5b. CHR:POS—chromosome and position of SNP according to GRCh37 human genome build; EA/RA—effective and reference allele; Gene—prioritized or nearest gene for a locus (Supplementary Table 6b); N—sample size; EAF—effective allele frequency; BETA (SE)—effect (in SD units) and standard error of effect; *P*—P-value; Top trait—glycan trait with the strongest association (the lowest *P*); Type of association—univariate or multivariate. Description of glycan traits is provided in Supplementary Table 3. Symbol ○ — the locus was previously discovered in immunoglobulin G N-glycome GWASs, as reviewed in ^25^.

| SNP info |  |  |  | Discovery |  |  |  |  |  |
| --- | --- | --- | --- | --- | --- | --- | --- | --- | --- |
| SNP | CHR:POS | EA/RA | Gene | N | EAF | BETA (SE) | *P* | Top trait | Type of association |
| rs12726286 | 1:93334379 | C/T | DIPK1A | 7540 | 73.52% | 0.022 (0.002) | 2.24E-21 | sialylation of antennary branches | multivariate |
| rs1329427 | 1:196704559 | C/T | CFH | 7540 | 57.63% | 0.016 (0.003) | 5.61E-10 | N-glycosylation | multivariate |
| rs1260326 | 2:27730940 | C/T | GCKR | 7081 | 57.51% | -0.168 (0.018) | 9.65E-22 | G3total | univariate |
| rs2470750 | 3:98690592 | A/T | ST3GAL6 | 6790 | 59.04% | 0.012 (0.002) | 7.23E-10 | high branching glycans | multivariate |
| rs3774964 | 4:103519487 | A/G | SLC39A8 | 7164 | 63.69% | 0.109 (0.018) | 1.49E-09 | G0n | univariate |
| rs7705720 | 5:95280033 | C/T | ELL2 ○ | 7540 | 20.03% | -0.133 (0.021) | 1.32E-10 | FG1S1/(FG1+FG1S1) | univariate |
| rs4543384 | 6:139629524 | C/T | TXLNB ○ | 6790 | 55.74% | 0.143 (0.018) | 9.67E-16 | FA2BG1n | univariate |
| rs7758383 | 6:143169723 | A/G | HIVEP2 ◯ | 7540 | 49.87% | 0.153 (0.017) | 3.55E-20 | FA2G2n | univariate |
| rs34166762 | 7:73018524 | C/T | MLXIPL | 7540 | 27.58% | -0.121 (0.019) | 1.09E-10 | A3G3S2 | univariate |
| rs7781265 | 7:150950940 | A/G | SMARCD3 ◯ | 7540 | 10.99% | 0.223 (0.027) | 4.75E-16 | M64 | univariate |
| rs4841133 | 8:9183664 | A/G | LOC157273 | 7540 | 8.37% | 0.013 (0.002) | 6.85E-10 | sialylation of antennary branches | multivariate |
| rs28601761 | 8:126500031 | C/G | TRIB1 | 6790 | 57.56% | -0.143 (0.018) | 7.10E-15 | G3Fa/G3total | univariate |
| rs582118 | 9:136145471 | A/G | ABO | 7540 | 65.12% | 0.023 (0.003) | 2.00E-17 | N-glycosylation | multivariate |
| rs174528 | 11:61543499 | C/T | FADS2 | 7540 | 35.83% | 0.013 (0.002) | 1.50E-09 | sialylation of antennary branches | multivariate |
| rs36020612 | 11:123344435 | C/T | GRAMD1B | 7319 | 79.96% | -0.169 (0.022) | 1.66E-14 | FBG2S1/<br>(FBG2+FBG2S1+FBG2S2) | univariate |
| rs11223982 | 11:134612702 | A/G | LINC02714 | 6280 | 12.47% | 0.256 (0.029) | 3.45E-19 | A4G4S3 | univariate |
| rs7161378 | 14:65450780 | C/T | MAX | 7540 | 24.74% | -0.180 (0.019) | 2.05E-20 | FG3/G3total | univariate |
| rs28929474 | 14:94844947 | C/T | SERPINA1 | 5135 | 97.71% | 0.437 (0.072) | 1.01E-09 | A2G2S2 | univariate |
| rs8046823 | 16:71400131 | A/G | CALB2 | 7540 | 51.53% | 0.021 (0.003) | 1.46E-16 | galactosylation of antennary<br>branches | multivariate |
| rs217184 | 16:72105965 | C/T | HP | 7540 | 19.90% | 0.211 (0.021) | 5.44E-23 | S3total | univariate |
| rs4500785 | 17:16848565 | C/G | TNFRSF13B ◯ | 7540 | 88.63% | -0.180 (0.027) | 1.33E-11 | FA2BG1 | univariate |
| rs2659007 | 17:79217478 | A/G | SLC38A10 ◯ | 7540 | 46.44% | 0.108 (0.017) | 4.07E-10 | FA2BG1n | univariate |
| rs11669860 | 19:19277296 | A/G | MEF2B ◯ | 7540 | 55.43% | -0.113 (0.017) | 7.14E-11 | FA2BG1n | univariate |
| rs2618588 | 20:17832658 | C/T | LOC107985440 ◯ | 7540 | 39.69% | 0.011 (0.002) | 2.02E-10 | core-fucosylation | multivariate |
| rs2834847 | 21:36588180 | A/C | RUNX1 ◯ | 7540 | 77.22% | 0.160 (0.020) | 2.58E-15 | FBn | univariate |

We performed an approximate conditional and joint analysis implemented in GCTA-COJO^45^ to identify conditionally independent association signals in the replicated loci on discovery GWAMA. We found evidence of multiple SNPs contributing independently to glycan level variation for nine loci (Supplementary Table 6a). Seven of these loci span glycosyltransferase genes, coding for enzymes directly involved in the biosynthesis of glycans. Two sentinel associations were observed in the loci containing fucosyltransferases *FUT8* and *FUT6*, sialyltransferases *ST6GAL1* and *ST3GAL4*, galactosyltransferase *B4GALT1,* glycuronyltranferase *B3GAT1*, and the acetylglucosaminyltransferase *MGAT5*. Beyond glycosyltransferase loci, the locus spanning the human leukocyte antigen (*HLA*) and the locus containing *HPR* gene showed secondary associations.

### SNP-based heritability and whole genome polygenic scores for 117 N-glycome traits

For 117 N-glycome traits we estimated SNP-based heritability using LD Score regression^44^. For 68 N-glycome traits SNP-based heritability was above zero at nominal P < 0.05, varying from 10.2% to 33.4% (19.8 ± 10.3%) (Supplementary Table 4с), which is on average 2.5x lower than the narrow-sense heritability of 37 N-glycome traits, estimated in a twins-based study – 50.6 ± 14.0%^46^.

For each of the 117 N-glycome traits, we created polygenic score (PGS) models based on the GWAMA of European-ancestry participants (N = 10,172) using the SBayesR method^47^. We tested the out-of-sample prediction accuracy of these models in the CEDAR dataset (N=187 participants of European ancestry). For 79 N-glycome traits in CEDAR samples, PGS models explained from 2.4% to 20.0% of the trait variance (FDR < 5%), allowing for calculation of genetically predicted glycan levels in large scale cohorts of European descent (e.g., UKBiobank). For the remaining 38 N-glycome traits the explained variance did not deviate significantly from zero (FDR > 5%). The out-of-sample prediction accuracy correlated significantly with the SNP-based heritability (R = 0.48, P = 4.05 x 10^-8^). The implementation of SBayesR models is detailed in Supplementary Table 7.

### Prioritization of causal genes for protein N-glycosylation

Identification of genes, rather than genetic loci, can help to find novel protein glycosylation regulators and suggest targets for intervention in glycome-related diseases. To prioritize the most likely effector genes, we employed a consensus-based prioritization approach selecting the gene with the highest unweighted sum of evidence from eight different predictors - based on a literature search of genes encoding known enzymes and regulators of N-glycan biosynthesis; genes causing congenital disorders of glycosylation; colocalization of glyQTLs with eQTLs and blood plasma pQTLs; annotation of putative causal variants affecting protein structure; enrichment of gene sets and tissue-specific expression; and prioritization of the nearest gene (see Methods). We prioritized the most likely effector gene for each locus by selecting the gene with the highest unweighted sum of evidence across all eight predictors^48^, provided a gene was supported by at least two predictors.

We prioritized candidate genes in 31 of the 40 glyQTLs (Supplementary Table 6b). The prioritized genes may regulate the protein N-glycosylation through several known general mechanisms: biosynthesis of N-glycans, abundance of N-glycoproteins in the blood, regulation of transcription in lymphoid and gastrointestinal tissues, and ion homeostasis in the endoplasmic reticulum and Golgi apparatus.

Among the 31 prioritized genes (**Fig. 2b**), we identified nine genes encoding glycosyltransferases (*MGAT5, ST6GAL1, B4GALT1, ABO, ST3GAL4, B3GAT1, FUT8, FUT6, MGAT3*); mutations in three are known to lead to congenital disorders of glycosylation (*B4GALT1, FUT8, SLC39A8*) and four genes have strong experimental support for being regulators of N-glycan biosynthesis genes (*HNF1A, IKZF1, RUNX3, SLC39A8*)^25^. The SMR/HEIDI approach indicated that total plasma N-glycosylation– associated variants in two loci possibly had pleiotropic effects on plasma levels of two blood proteins (HPT, CAFH) (Supplementary Table 6g) and transcription of 10 genes in different tissues (Supplementary Table 6f). In 12 genes, associated variants were either coding or were in strong LD with the variants coding for potentially deleterious amino acid changes (annotated by Variant Effect Predictor, VEP^49^), and in 5 genes - pathogenic amino acid changes (predicted by FATHMM XF^50^ and FATHMM InDel^51^). The DEPICT gene prioritization tool^31^ provided evidence of prioritization for 19 genes in 18 loci at FDR < 0.2 (Supplementary Table 6h).

**Figure 2:**
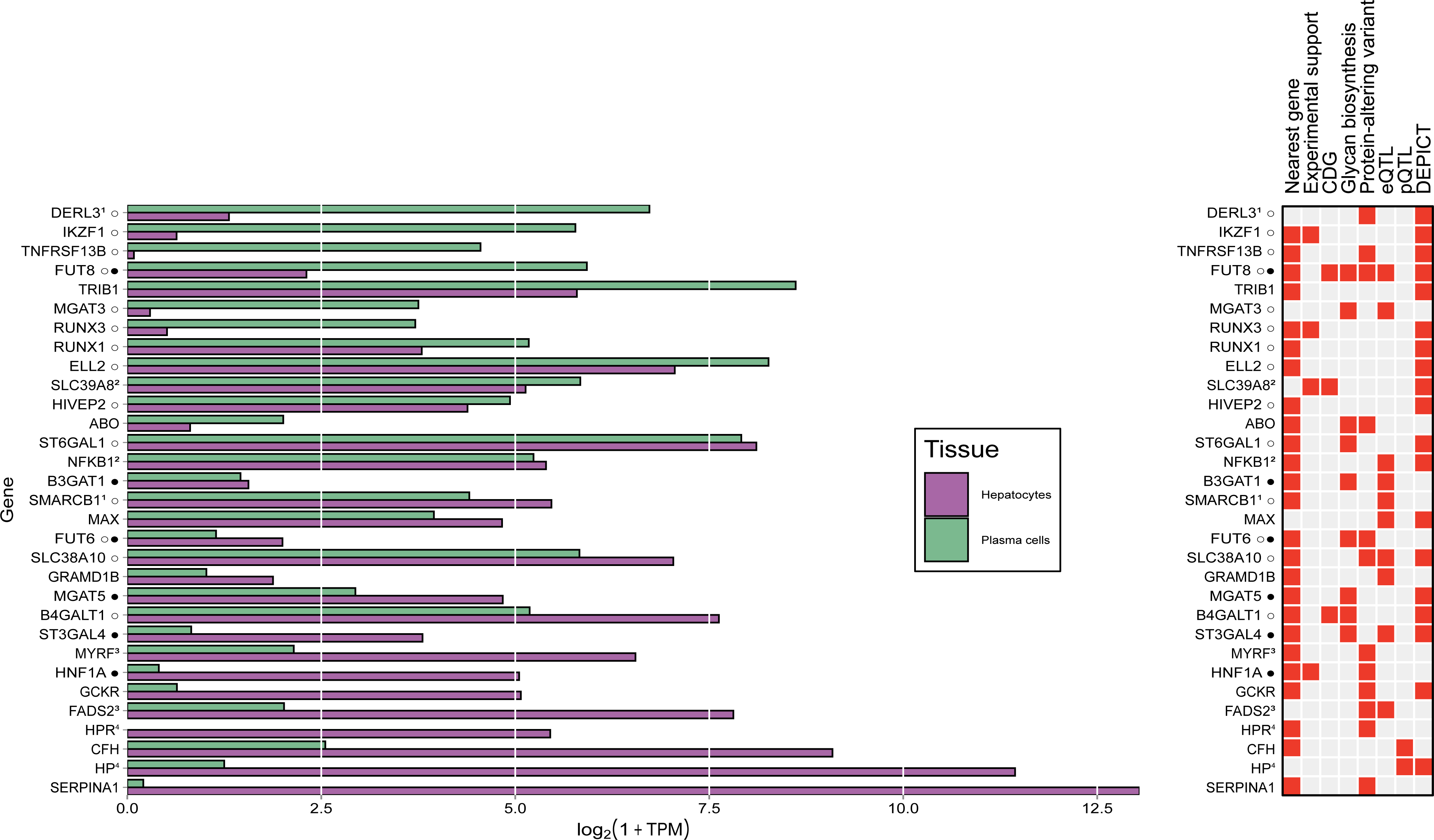
Сandidate genes. (a) Gene expression of the candidate genes in two relevant cell types - hepatocytes and plasma cells. Expression levels are represented as the median logarithm of transcripts per million. The data for hepatocytes (N = 513) and plasma cells (N = 53) samples were obtained from the ARCHS4 portal ^88^. (b) Predictors indicating the 32 candidate genes. Gene order corresponds to (a). The identical superscripts denote candidate genes inside one locus. Full details of the gene prioritization are presented in Supplementary Table 6b. Symbol ○ next to the candidate gene indicates that the locus was previously discovered in immunoglobulin G N-Glycome GWASs; Symbol ● — the locus was previously discovered in transferrin N-Glycome GWAS, as reviewed in ^25^.

Because not all the glyQTLs were colocalized with a cis-eQTL, cis-pQTL or lay in proximity to biologically relevant genes, we also utilized the nearest protein-coding genes as an independent predictor. This approach was chosen due to the tendency of the nearest protein-coding genes to enrich for molecular QTLs^52^.

In the following discussion, we focus on thirteen novel candidate genes that were not identified before in GWASs of human protein N-glycosylation; for the latter, we refer a reader to previous works and published reviews^25^. We prioritized four genes associated with lipid metabolism regulation - *GCKR*, *FADS2*, *TRIB1*, and *GRAMD1B* which, to our knowledge, is the first time protein N-glycosylation has been linked to genes involved in lipid metabolism and its regulation; four genes encoding N-glycoproteins having anti-inflammatory function - *HP, HRP, SERPINA1* and *CFH*; the gene *SCL39A8* encoding a zinc transporter; three genes encoding transcription factors - *MAX*, *NFKB1*, *MYRF*; and a glycosyltransferase gene *ABO*, which determines an individual’s ABO blood type.

The Supplementary Note provides an in-depth account of the details of thirteen newly prioritized genes. The other genes that have been previously prioritized elsewhere are described in Timoshchuk et al.^25^.

### Tissue-specific regulation of plasma protein N-glycosylation

Lymphoid tissue, specifically plasma cells that produce antibodies, and liver, specifically hepatocytes, contribute the majority of glycoproteins present in human blood^2,25^ and are thus the primary drivers of N-glycosylation of plasma proteome. However, the N-glycosylation machinery in these two cell types varies, leading to distinct spectra of glycans attached to proteins produced in these two tissues^2,46^. Many glycosylation-associated genes prioritized in this study are expressed in plasma cells, or hepatocytes, or both (**Fig. 2a**).

To gain a deeper understanding of the tissue-specific regulation of glyco-genes, we constructed a gene network for N-glycosylation regulation. This network comprised 32 loci that were replicated in the univariate association analysis, and 117 N-glycome traits as vertexes, with significant associations between them represented as edges (**Fig. 1b**). The resulting network revealed two major subnetworks, wherein candidate genes and glycan traits were clustered. The first subnetwork was primarily associated with blood plasma N-glycans typically produced in the liver, and included 13 loci (*ATF6B, B3GAT1, CHST2, FUT6, HNF1A, HP, LINC02714, MAX, MGAT5, MLXIPL, SERPINA1, ST3GAL4, TRIB1*). The second subnetwork was related to blood plasma N-glycans typically attached to immunoglobulins and consisted of 14 loci (*B4GALT1, ELL2, HIVEP2, IKZF1, MEF2B, MGAT3, RUNX1, SLC38A10, SLC39A8, SMARCB1, SMARCD3, TNFRSF13B, TMEM121, TXLNB*). According to classification of Clerc et al.^2^, genetic variation in five loci (containing *FUT8, GCKR, GRAMD1B, RUNX3,* and *ST6GAL1*) had an impact on plasma N-glycans attached to both immunoglobulins and liver-secreted proteins. Most of these five loci exhibited strong bias towards N-glycans known to be preferentially expressed on proteins produced in one of the tissues, i.e., *GRAMD1B* was associated with 8 N-glycans, of which 7 were typical for liver proteins; *GCKR* – with 9, of which only one was typical for immunoglobulins; *RUNX3* and *ST6GAL1* were preferentially associated with N-glycans typically attached to immunoglobulins (32/35 and 43/44 glycans, respectively). It should be noted that the classification of Clerc et al.^2^ was compiled based on a large body of literature data, and we cannot exclude occasional misclassification.

To gain insight into the spectrum of glycans that were associated to the 8 loci that were replicated in multivariate association analysis, we considered significant (at p < 0.01/36) association of the partial regression coefficients in the multivariate analysis of trait set “N-glycosylation” (36 traits) (Supplementary Table 5e). In this analysis, *DIPK1A*, *FADS2*, and *CALB2* loci associated with N-glycans typical for liver-secreted proteins; *LOC107985440* – with N-glycans typically observed on immunoglobulins IgG. Results for *ST3GAL6* and *LOC157273* were inconclusive, although the former was associated with the multivariate trait “high branching N-glycans”; such glycans are typical for liver-secreted proteins.

Three loci showed clear effect on N-glycans found on both liver-secreted glycoproteins and immunoglobulins: *FUT8* (significant effect on 9 N-glycans typically attached to liver-secreted proteins and 3 typically attached to immunoglobulins); *CFH* (2 and 2, respectively) and *ABO* (also 2 and 2).

TF and IgG are two proteins secreted by hepatocytes and plasma cells, respectively, and GWASs of their N-glycosylation shed light on the genetic control of protein N-glycosylation in the corresponding tissues^42^. To gain further insights into the mechanism of association and to support the tissue-specificity of the loci, we conducted a colocalization analysis of total plasma, IgG and TF glyQTLs using the SMR-θ method^53^. The analysis was restricted to the loci that were previously implicated in TF^42^ N-glycome or IgG^40^ N-glycome GWASes, and reached genome-wide significance in univariate association analysis in this study. Excluding *HLA*, this selection resulted in 21 loci, of which 15 were significant in previous IgG N-glycome GWAS only, four were only significant in the TF N-glycome GWAS, and two (*FUT6* and *FUT8*) were significant in both (Supplementary Table 5d)^25^. For specific locus, we colocalized signals of genetic association for traits that have reached genome-wide significance in that locus.

The results of colocalization analysis are presented in Supplementary Figures 3. If regional genetic associations of a plasma N-glycome trait colocalized (|θ|>0.7) with genetic associations of an IgG N-glycome trait, we considered this as evidence that the locus is expressing its effect on plasma N-glycome through its effect on IgG N-glycosylation, acting in antibody-producing cells. Similarly, colocalization with genetic association signal for TF N-glycome was taken as an indication that the locus may exhibit its action via effect of TF N-glycome, acting in liver. The analysis suggested that the *ELL2, TXLNB, HIVEP2, IKZF1, SMARCD3, TMEM121, SLC38A10, MEF2B, ATF6B, RUNX1, RUNX3, SMARCB1, MGAT3, ST6GAL1, B4GALT1* loci regulate N-glycosylation of IgG while *MGAT5, ST3GAL4, B3GAT1, HNF1A* loci regulate N-glycosylation of TF. The *FUT8 and FUT6* act as regulators of both glycoproteins. An interesting case of pleiotropy was observed in the *FUT8* locus. The colocalization signal in *FUT8* split into two distinct clusters (Supplementary Fig. 3, page 120), one of which was dominated by N-glycans predominantly presented on proteins produced in the liver, while the other was almost exclusively presented by these on immunoglobulins. To support the hypothesis of two distinct tissue-specific genetic mechanisms in the locus, we combined traits from the two clusters into single traits using MANOVA approach^28^ and performed a colocalization analysis between the two constructed linear combinations using the SMR-θ method^53^ and R Coloc package^54^. We found strong evidence *against* colocalization of N-glycans presented on liver-specific proteins and these on immunoglobulins in this genomic region (PP.H3 = 100%, where PP.H3 is the posterior probability that there are two distinct causal variants contributing to trait variation, θ = 4 x 10^-6^). This supports the hypothesis that genetic regulation of *FUT8* in the liver and antibody-producing cells follows two distinct mechanisms, as previously suggested by Landini and colleagues^42^.

Thus, the colocalization analysis confirms different tissue-specific mechanisms of genetic regulation for each locus. With an exception of the *ATF6B* and *FUT6* loci, the results from colocalization are largely consistent with the classification based on gene-N-glycan association network.

Our findings demonstrate that the genetic regulation of protein N-glycosylation is highly tissue-specific. Even glycosyltransferases such as *FUT8*, *FUT6, MGAT3*, *ST6GAL1* and *B4GALT1*, being expressed in antibody-producing cells and hepatocytes (Fig. 2a) and known to participate in the N-glycan biosynthesis in both tissues, display pronounced tissue-specific genetic regulation that is not shared between different tissues.

### PheWAS highlights loci associated with an extensive number of diseases and quantitative traits

In this study, we examined the pleiotropic effects of 40 replicated glyQTLs on over a thousand diseases and quantitative traits endpoints (as listed in Supplementary Table 8a) using the SMR/HEIDI method. Our analysis revealed a total of 1,214 significant associations, of which 781 demonstrated a non-rejection of the pleiotropy hypothesis by the HEIDI test. The identified pleiotropic associations encompassed a wide range of phenotypes, including type 2 and type 1 diabetes, blood glucose levels, coronary artery disease, cholesterol levels, bipolar disorder, schizophrenia, gout, various oncological diseases, metabolomic and anthropometric traits, lifestyle and diet-related traits, general life history and overall health status, among others (as depicted in **Fig. 3a, 3b** and Supplementary Table 8b). Additionally, *TRIB1* and *GCKR* showed colocalization with metabolic dysfunction-associated steatotic liver disease (*TRIB1*: P_SMR_ = 4.58 x 10^-8^, P_HEIDI_ = 0.03 (possibly shared); *GCKR*: P_SMR_ = 3.26 x 10^-8^, P_HEIDI_ = 0.73 (likely shared)).

**Figure 3:**
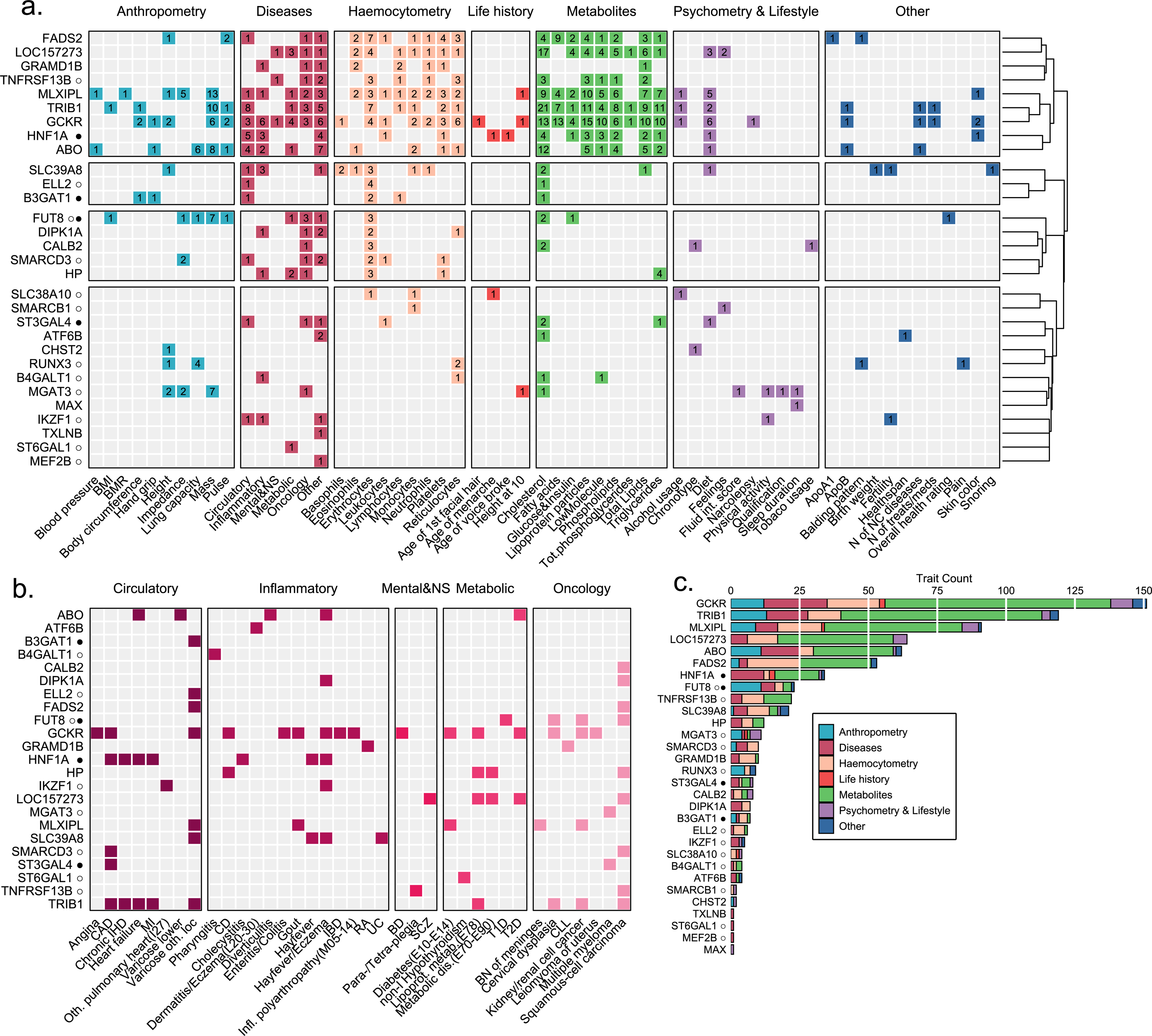
Phenome-wide colocalization results for significant and replicated loci. (a) Heatmap of traits with pleiotropic associations (PadjSMR < 0.05 and PHEIDI >= 0.05). The row order is determined by clustering on the set of significantly colocalized traits (Jaccard similarity index and Ward’s linkage). For the sake of readability similar traits are grouped into broader categories, e.g., such traits as fat-free mass of left leg, trunk fat mass are combined under the name “Body mass”. The numbers in the cells represent the number of pleiotropic associations, grouped under a common name, confirmed for a given locus. Color does not carry semantic load. Heatmap providing details of pleiotropic associations with diseases from (a). Color does not carry semantic load. (b) Count of traits for each locus with pleiotropic associations. Symbol ○ next to the candidate gene indicates that the locus was previously discovered in immunoglobulin G N-Glycome GWASs; Symbol ● — the locus was previously discovered in transferrin N-Glycome GWAS, as reviewed in ^25^. Abbreviations: BMI: body mass index; BN: benign neoplasm; CLL: chronic leukocytic leukemia; T1D: type 1 diabetes; T2D: type 2 diabetes; BD: bipolar disorder; BP: blood pressure; MI: myocardial infarction; CAD: coronary artery’s disease; CD: Crohn disease; IBD: Inflammatory bowel disease; IHD: Ischemic heart disease; NC: non-cancer, non-I: non-iodine dependent; RA: rheumatoid arthritis; SCZ: schizophrenia; UC: ulcerative colitis.

Hierarchical clustering based on sets of colocalized traits allowed us to differentiate four distinct groups of loci (**Fig. 3a**, right panel). The first cluster (**Fig. 3a**, topmost cluster) was characterized by a high number of metabolic colocalizations and encompassed eight of the ten most pleiotropic loci (**Fig. 3c**). It was also found to be colocalized with a diverse range of other phenotypes, including disease and anthropometric traits. Of particular note, this cluster comprised loci with prioritized lipid metabolism genes, namely, *GCKR, TRIB1, FADS2* and previously known *HNF1A*.

Furthermore, it encompassed the locus containing the *MLXIPL* gene, a transcriptional factor that induces liver glycolysis and lipogenesis, as well as *ABO*, which is known to be associated with stroke^55^, metabolic dysfunction-associated steatotic liver disease and levels of lipids^56,57^.

### Association between PGS for plasma N-glycosylation traits and ICD-10 diseases

We analyzed associations between polygenic scores (PGS) for the 117 plasma N-glycosylation traits and 167 diseases classified according to International Classification of Diseases (ICD)-10 in individuals of European ancestry from the UK Biobank cohort (N = 374,303) (Supplementary Note). The analysis revealed 14 diseases associated with PGS for at least one plasma N-glycome trait and PGS for 35 plasma N-glycome traits associated with at least one disease at the designated significance threshold of p < 1.07 x 10^-5^ (**Fig. 4**, Supplementary Table 9). Full results and overview of the analysis are provided in the Supplementary Note.

**Figure 4:**
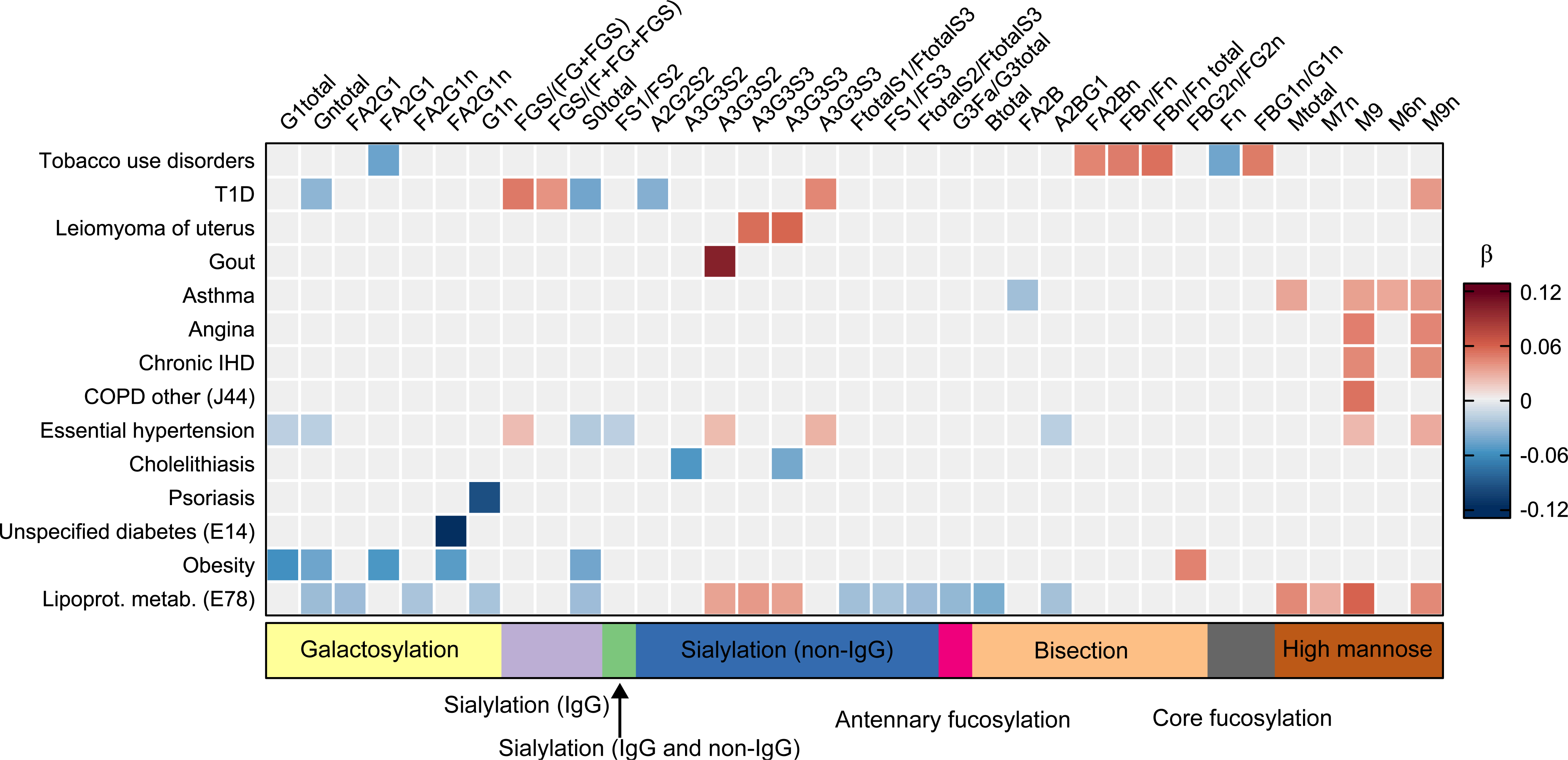
Significant associations between PGS for plasma N-glycosylation traits and ICD-10 diseases. Heatmap of associations between PGS for plasma N-glycosylation traits and ICD-10 diseases that were statistically significant at the designated significance threshold of 1.07 x 10-5. Every column represents an N-glycosylation trait, every row – an ICD-10 disease. The colour of the cell represents the effects estimated in the logistic regression of the disease phenotype incidence on the PGS for each glycan trait – blue hues represent negative effects, red hues – positive, and the intensity of the colour represents the absolute value of the effect (larger values are shown in darker hues). Grey cells represent the non-significant associations. The bands under the heatmap depict the groups of N-glycosylation traits that are related to certain structural features of the N-glycans. T1D – type 1 diabetes.

Notably, we observed positive associations between PGS for traits, related to the increased levels of high-mannose glycans, especially to those of containing nine mannsoe residuals (M9), with cardiovascular disease phenotypes such as essential hypertension, ischemic heart disease, angina, hyperlipidaemias, as well as type 1 diabetes and asthma (**Fig. 4**).

We found negative associations for the PGS for S0 total (percentage of neutral N-glycan structures, i.e. N-glycans without sialic acid, in total plasma N-glycans) with primary hypertension, lipidaemias, obesity and non-insulin dependent diabetes (**Fig. 4**). Moreover, PGS for the N-glycome traits describing the abundances of galactosylated structures were negatively associated with obesity, lipoprotein metabolism disorders, primary hypertension and type 2 diabetes (**Fig. 4**).

### Bidirectional Mendelian Randomization analysis of causal effects between plasma N-glycosylation traits and ICD-10 diseases

For the statistically significantly associated pairs of disease-plasma N-glycome traits in UK Biobank we conducted bidirectional two-sample Mendelian randomization (MR) analysis to investigate the direction of effects.

Using the disease phenotype as exposure, we conducted a two-sample MR analysis and revealed statistically significant positive causal effect of disorders of lipoprotein metabolism on M9 (N-glycan with nine mannose residuals) and on Mtotal (the percentage of high-mannose structures in total plasma glycans) (**Table 2**, Supplementary Table 10b, Supplementary Fig. 6a-d, 7a-d). The direction of the causal effect corresponded to the signs of the beta of association between disorders of lipoprotein metabolism and other lipidaemias (E78) and PGS for M9 and Mtotal. Sensitivity analyses, including a two-sample MR after removal of pleiotropic IVs, as well confirmed the observed causal effects of lipidaemias (E78) on M9 and Mtotal (Supplementary Fig. 8a-d, 9a-d; Supplementary Tables 11a-c, 13a-c, Supplementary Note).

**Table 2:** Causal associations between plasma N-glycosylation traits and ICD-10 diseases discovered in this study. Full results of the analysis are provided in Supplementary Tables 10a, 10b. Exposure – upstream trait in the analysis (cause); Outcome – downstream trait in the analysis; IV-instrumental variable (independent SNPs associated with upstream trait); MR – Mendelian randomization; Causal BETA (SE) – causal effect of the exposure on the outcome (in log OR units of the disease unit per SD units of glycan trait if glycan trait was an exposure, or in SD units of glycan trait per log OR unit of the disease phenotype if disease phenotype was an exposure) with its standard error; *P* - P-value of the MR analysis.

| Exposure | Outcome | MR method | Number of IVs | Causal BETA (SE) | *P* |
| --- | --- | --- | --- | --- | --- |
| PGP64 - The percentage of M6 in total neutral plasma glycans (GPn) | J45 - Asthma | Weighted median | 3 | 0.012 (0,0036) | 9.78E-04 |
| PGP69 - The percentage of M9 in total neutral plasma glycans (GPn) | E78 - Disorders of lipoprotein metabolism and other lipidaemias | Inverse variance weighted | 2 | 0.013 (0,0035) | 1.25E-04 |
| E78 - Disorders of lipoprotein metabolism and other lipidaemias | PGP18 - The percentage of M9 | Inverse variance weighted | 27 | 4.09 (1,1772) | 5.16E-04 |
| E78 - Disorders of lipoprotein metabolism and other lipidaemias | PGP107 - The percentage of high-mannose structures in total plasma glycans | Weighted median | 27 | 3.01 (0,8756) | 5.87E-04 |

Using plasma N-glycome trait as exposure we performed MR analysis in the opposite direction, discovering a statistically significant positive causal effect of M6n, percentage of M6 in total neutral plasma glycans on asthma and a positive effect of M9n, percentage of M9 in total neutral plasma glycans, on disorders of lipoprotein metabolism and other lipidaemias (**Table 2**, Supplementary Table 10a, Supplementary Fig. 4a-d; 5a-c). In both cases, the direction of the effect was concordant with the direction of association between corresponding pairs of disease and PGS. Sensitivity analyses also confirmed the observed causal effect (Supplementary Tables 11a-c). Since the number of available IVs for both M6n and M9n was not sufficient for the analysis of pleiotropy among the IVs using MR-PRESSO^58^, as an additional sensitivity analysis for the effects of M6n on asthma and M9n on lipidaemia we performed colocalization (SMR-HEIDI) analysis for the loci tagged by the genetic variants used as IVs in these cases (Supplementary Table 12). SMR-HEIDI analysis provided evidence for one shared causal variant (rs144126567, p_SMR_ = 0.003; p_HEIDI_ = 0.30) for M6n and asthma but did not find any proof for existence of shared genetic variants influencing M9n and lipidaemia (Supplementary Table 12). Therefore, we report an observed positive effect of M6n on asthma, while the presence of a positive effect of M9n on lipidaemia remains inconclusive. Full results of MR analysis are presented in Supplementary Tables 10a-b.

## Discussion

Here, we reported 40 quantitative trait loci (glyQTLs) discovered in the GWAS of 138 blood plasma N-glycome traits, resulting in a more than two-fold expansion of loci affecting N-glycosylation of blood plasma proteins. The integration of these findings with genetic information related to human diseases and other complex phenotypes allowed us to show for the first time that genes involved in liver function are linked to the human blood plasma protein N-glycosylation.

A subset of newly prioritized genes allows us to postulate a link between genetic regulation of metabolic and liver diseases and blood plasma protein N-glycosylation.

Specifically, common genetic variation in the loci near *GCKR and TRIB1* is known to predispose to metabolic dysfunction-associated steatotic liver disease (MASLD)^59^. Moreover, genetic association signal for MASLD in these loci are colocalized with the corresponding glyQTLs. In the gene *SERPINA1*, rare Mendelian mutations lead to alpha-1 antitrypsin (AAT) deficiency, with liver disease as part of the phenotype. Common variation in this region associates with chronic elevation of alanine aminotransferase (cALT) levels^60^, a proxy phenotype for MASLD.

Although on phenotypic level, the changes of total and liver secreted protein-specific N-glycosylation in liver disease are well-known^61–63^, we demonstrate, for the first time, that specific genes are associated with both N-glycosylation and liver disease, offering a starting point for genetically-guided investigation of the functional mechanisms of this phenotypic association.

Somewhat superficially, we may reason that liver disease is characterized by hepatocyte injury and endoplasmic reticulum stress^64^, which is strongly associated with changes in glycosylation. A less direct mechanism could be inflammation, that is a hallmark of liver disease, with proinflammatory cytokines shown to alter the substrate synthesis pathways as well as the expression of glycosyltransferases required for the biosynthesis of N-glycans^65^. Thus, changes in N-glycosylation observed in blood plasma may be at least partly explained by altered N-glycosylation of hepatocyte-secreted proteins. Consistently with this hypothesis, we demonstrate that common genetic variation in the three loci known to be associated with liver disease is associated with variation in abundance of N-glycans, typically attached to the liver-secreted proteins.

Other notable, partly overlapping, subset of liver-expressed genes newly implicated in plasma protein N-glycosylation encodes anti-inflammatory proteins -- haptoglobin (HP, *HP*), complement factor H (CFAH, *CFH*), and alpha-1-antitrypsin (AAT, *SERPINA1*). The glyQTLs located at *HP* and *CFH* are colocalized with the corresponding pQTLs. At least two mechanisms may be suggested to explain such colocalization. Genetic variants in these loci may affect the composition of blood N-glycosylation through changes in the abundance of glycans preferentially bond to HP and CFAH by regulating the level of these glycoproteins in the blood. Alternatively, the genetic variation may change glycosylation of these proteins, which, in turn, could change the affinity of the binding of the specific probes used by the SomaLogic assays. While we did not observe colocalization between *SERPINA1* glyQTL and the AAT pQTL, this may be a false negative, potentially explained by the low frequency of the *SERPINA1* lead variant (rs28929474). Nonetheless, the rs28929474 is associated with the level of glycoprotein acetyl, a mixture of N-glycoproteins, predominantly alpha-1 acid glycoprotein, haptoglobin, and alpha 1 antitrypsin^66^.

While variation at *HP* and *SERPINA1* loci is associated with changes in N-glycans typically attached to liver proteins, *CFH* locus appears to also affect N-glycans typically attached to immunoglobulins. While *CFH* does express at some level in plasma cells (**Fig. 2a**), we speculate that perhaps a more likely mechanism is that genetic variation in *CFH* affects its expression liver, and changes in N-glycans attached to immunoglobulins occur through a systemic mechanism, i.e., regulation of inflammation. Consistent with this hypothesis is the known association of the *CFH* locus with IgA nephropathy^67^, as well as indications that in mice, that CFH modulates splenic B cell development and limits autoantibody production^68^.

Our results suggest that genetic regulation of plasma protein N-glycosylation predominantly occurs in lymphoid and liver tissue and exhibits strong tissue specificity. Integration of evidence from transcriptomics and N-glycomics suggests that molecular expression of genetic variation in the majority of glyQTLs is restricted to one tissue; the effects of this variation on N-glycans in blood plasma occur either through changes of N-glycosylation of the proteins secreted by the tissue, or through systemic mechanisms. Of note, all of N-glycosyltransferase genes that express on RNA, protein, and glycan levels in both tissues, are genetically regulated in only one of them (*B4GALT1, ST6GAL1, MGAT3*), or exhibit different glyQTLs in different tissues (*FUT8, FUT6)*. Even thus, while a number of glycosyltransferases are expressed both in liver and lymphoid tissues, we provide evidence that the genetic variation regulating N-glycosylation in each of the two tissues is unique and does not overlap at the available resolution of the analysis. To the best of our knowledge, this is the first study to analyze and reveal the strong tissue-specificity of the genetic regulation of population variability of human protein N-glycosylation.

Further studies of the genetic regulation of N-glycosylation of individual proteins rather than bulk N-glycome will lead to the discovery of novel glyQTLs, which we cannot observe now due to a lack of power or noise in bulk N-glycome. Quantification of the N-glycome of purified proteins like IgG^40^, TF^42^, IgA^69,70^, and apolipoprotein CIII^71^, and other proteins, will be highly relevant to understanding the etiology of such disease, as rheumatoid arthritis, hepatocellular carcinoma, IgA nephropathy, endocarditis^7^. Alternatively, development and application of computational deconvolution approaches may be similar to those applied for bulk RNA-Seq^72^.

In conclusion, our study offers insight into the genetic factors influencing blood plasma N-glycome, and, for the first time, establishes a genetic link between N-glycosylation, liver disease, and anti-inflammatory proteins. The identification of novel genes associated with metabolic and liver disease and N-glycosylation contributes to deeper understanding of shared biological mechanisms and will facilitate future biomarker discovery and interpretation.

## Methods

We conducted a multicenter study using data from seven studies – TwinsUK (N = 3,918), EPIC-Potsdam (N = 2,192), PainOmics (N = 1,873), SOCCS (N = 1,742), SABRE (N = 544), QMDiab (N = 325), and CEDAR (N = 170) with a total sample size N = 10,764. Local research ethics committees approved all studies, and all participants gave written informed consent. The detailed description of the cohorts is shown in Supplementary Table 1 and Supplementary Notes.

### Glycome measurement and phenotype processing

Plasma N-glycome quantification of samples from all but SOCCS studies was performed at Genos Ltd using the protocol published previously^73^. Briefly, plasma N-glycans were enzymatically released from proteins by PNGase F, fluorescently labeled with 2-aminobenzamide and cleaned-up. Fluorescently labeled and purified N-glycans were separated by HILIC on a Waters BEH Glycan chromatography column. The fluorescence detector was set with excitation and emission wavelengths of 250 nm and 428 nm, respectively. Plasma N-glycome quantification for SOCCS samples was done at NIBRT by applying the same protocol with the only difference in the excitation wavelength (330 nm instead of 250 nm). Glycan peaks (GPs) – quantitative measurements of glycan levels – were defined by manual integration of intensity peaks in the chromatograms or were defined by automatic integration. The number of defined GPs varied among studies from 36 to 42, therefore to conduct multi-center association analysis followed by meta-analysis, we harmonized the set of GPs by applying a recently published protocol^34^ to a harmonized set of 36 GPs. To reduce experimental variation in glycan measurements, before genetic studies, raw glycan data were probabilistic median quotient normalized^74,75^ and batch corrected centrally by the phenotype provider (Genos Ltd). More detailed information on glycan preprocessing can be found in the Supplementary Note. From the 36 directly measured glycan traits, 81 derived traits were calculated (Supplementary Table 3a). These derived traits average glycosylation features such as branching, galactosylation, and sialylation, etc, across different individual glycan structures and, consequently, they may be more closely related to individual enzymatic activity and underlying genetic polymorphism.

### Discovery and replication genetic association analysis

#### Single trait association analysis

Discovery genome-wide association studies were performed in (sub) cohorts of European descent: TwinsUK (N = 2,739), EPIC-Potsdam (N = 2,192), PainOmics (N = 1,873), SOCCS (controls, N = 459) and SABRE (N = 277) with a combined sample size of 7,540 (Supplementary Table 1b). Prior to GWAS, the total plasma N-glycome traits were adjusted for sex and age, and the residuals were quantile transformed to normal distribution. The genetic association analysis in each cohort was conducted using a similar protocol. We assumed an additive model of genetic effects. GWAS were based on the genotypes imputed from Haplotype Reference Consortium Results^76^ or 1000 Genomes project^77^. Results of GWAS in each discovery cohort passed a strict quality control procedure followed by fixed-effects inverse-variance weighted meta-analysis. After quality control, 8.8 M SNPs were used for the downstream analysis.

To define genome-wide significant glyQTLs, we used conventional genome-wide significance threshold, Bonferroni corrected by 28 independent glycan traits (P < 1.79 x 10^-9^) as suggested before^78^. We considered SNPs located in the same locus if they were located within 250 Kb from the leading SNP (the SNP with lowest *P*). Only the SNPs and the traits with lowest *P* are reported (leading SNP-trait pairs) in the **Table 1**. The detailed procedure of locus definition is described in Supplementary Note.

Replication GWAS were performed using (sub) cohorts of samples with European descent: SOCCS (colorectal cancer cases, N = 1,283), TwinsUK (N = 1,179), CEDAR (N = 170); South Asian descent: SABRE (N = 267) and Arabian, Indian, Filipino descent: QMDiab (N = 325) (Supplementary Table 1b). Results of GWAS in each discovery cohort passed a strict quality control procedure followed by fixed-effects inverse-variance weighted meta-analysis. For replication of novel glyQTL, found at the discovery step, we used the leading SNP-trait pair that showed the most significant association. The replication threshold was set as P < 0.05/28 = 0.00178, where 28 is the number of replicated loci. Moreover, we checked whether the sign of estimated effect was concordant between discovery and replication studies.

#### Identification of secondary associations in glyQTLs

To identify secondary association signals at glyQTL in univariate analysis and capture the overall contribution to phenotypic variation, we performed conditional analysis using GCTA-COJO software, version 1.93.2beta^45^. This method uses summary-level statistics from a discovery meta-analysis and LD corrections between SNPs estimated from a reference sample for implementing a stepwise selection procedure including a series of conditional and joint regression analyses in which the SNP with the strongest association in the region is added to the regression model until no additional SNPs reach genome-wide significance. We used 1,429 unrelated individuals with European descent from SABRE cohort as reference samples for LD calculation. We used P < 1.79 x 10^-9^ as a genome-wide significance level and a default window setting to identify lead associations (Supplementary Table 6a).

#### Multi-trait association analysis

To gain additional power of glyQTL detection, we performed a multivariate GWAS of total plasma N-glycome. It has been previously demonstrated that multivariate genetic association analysis of N-glycome, that is, a joint analysis of multiple N-glycome traits, has higher power for loci detection than a regression model under which glycome traits are analyzed independently of each other^28,41^.

For discovery and replication analyses, we used discovery and replication GWAMA summary statistics, obtained in single-trait analysis. The discovery multivariate analysis was performed using the MANOVA-based method, adopted for analysis of a group of single-trait GWAS summary statistics (details are in Supplementary Note Discovery multivariate analysis)^28^. Discovery analysis was performed using the MultiABEL R package. Due to method requirements, we filtered out SNPs with sample size lower than 6,790 (which is 90% of 7,540 samples). The statistical significance threshold for multivariate analysis was set at P < 5 x 10^-8^ /21, where 21 is the number of multivariate traits described in Supplementary Table 3b and Supplementary Note. GlyQTLs with significant association were defined in the same way as for single-trait discovery.

For replication of multi-trait associated glyQTLs we used a complex four-step replication strategy as proposed by Ning et al.^29^, which consists of the following steps: MANOVA, Phenotype Score, Pearson correlation method and Kendall correlation method. In the first step (MANOVA) we straightforwardly checked whether the locus is significantly associated with the multivariate trait in the replication cohort using the same test as in the discovery stage. The replication threshold was set as 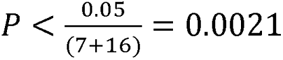, where 7 is the number of previously identified but not replicated loci and 16 is the number of novel loci. Then we checked whether the effect direction is consistent between the two cohorts, using the phenotype score approach^29^. Next, we evaluated the concordance of multivariate effect between two samples using Pearson and Kendall’s correlation coefficients. We considered an association of the locus replicated if it had successfully passed MANOVA and phenotype score steps of replication. The multivariate effect of the locus replicated if it additionally had passed both Pearson’s and Kendall’s correlation steps of replication (Supplementary Note, Supplementary Table 5b, and Supplementary Fig. 2). Phenotype score-based replication was performed as in Shadrina et al^41^. For each lead pair of SNP and trait group phenotype, we extracted coefficients of the linear combination of genotype onto multiple atomic phenotypes, estimated for discovery cohort. We used them to construct the corresponding trait group phenotypes for further testing of an association between the lead SNPs and the derived linear combinations (see Supplementary Note for details). A locus was replicated if the association of the SNP with the constructed linear combination had the same direction of effect as in the discovery cohort and passed the threshold of P < 0.0021.

To evaluate the similarity between estimates of multivariate genetic effects from discovery and replication cohorts across multiple traits, we used an MC-based approach implemented in MultiABEL package (MV.cor.test() function)^28^. For both Pearson’s and Kendall’s correlation coefficients, we considered a multivariate effect for a specific SNP replicated if the 95% confidence intervals didn’t include zero.

### SNP-based heritability and polygenic scores

SNP-based heritability was estimated using the LD Score regression software^44^ embedded in the GWAS-MAP platform^79^. We used pre-computed LD scores that were calculated from the European-ancestry samples in the 1000 Genomes Project. Only the 1,176,189 HapMap3 SNPs were included with a MAF_>_0.05. For the purpose of heritability estimation and further post-GWAS analyses, we generated GWAMA summary statistics for the samples of European descent with N = 10,172. We used GWAMA summary statistics for the analysis in order to use the largest data set with homogeneous ancestry.

SBayesR method reweights the effect of each variant according to the marginal estimate of its effect size, statistical strength of association, the degree of correlation between the variant and other variants nearby, and tuning parameters. This method requires a compatible LD matrix file computed using individual-level data from a reference population. For these analyses, we used publicly available shrunk sparse GCTB LD matrix including 1.1 million HapMap3 variants and computed from a random set of 50,000 individuals of European ancestry from the UKB data set^47,80^. SBayesR (gctb_2.03) was run for each chromosome separately, and with the default parameters except for the number of iterations (N = 5,000) and options for the stability of the algorithm (Supplementary Table 7). The prediction accuracy was defined as the proportion of the variance of a phenotype that is explained by PGS values (R2). To calculate PGS based on the PGS model, we used PLINK2 software^81^, where PGS values were calculated as a weighted sum of allele counts. Out-of-sample prediction accuracy was evaluated using samples from the CEDAR cohort that were not used for discovery or replication.

### Prioritization of candidate genes in found loci

For the purpose of post-GWAS analyses, we generated GWAMA summary statistics for the samples of European descent (N = 10,172). GWAMA summary statistics passed the same QC procedure as discovery and replication GWAMA. We applied an ensemble of methods to prioritize plausible candidate genes in the loci with found and replicated glyQTL (32 in univariate and 8 in multivariate analysis). We applied eight approaches to prioritize the most likely effector genes: (1) prioritization of the nearest gene; (2) prioritization of genes with known role in biosynthesis of N-glycans; (3) genes of congenital disorders of glycosylation; (4) genes with direct experimental support for regulation of protein N-glycosylation; (5) prioritization of genes containing variants in strong LD (r^2^ ≥ 0.8) with the lead variant, which are protein truncating variants (annotated by Variant Effect Predictor, VEP^49^) or predicted to be damaging by FATHMM XF^50^, FATHMM InDel^51^; (6) prioritization of genes whose eQTL and/or (7) pQTL are colocalized with glyQTL; (8) prioritization of genes based on the gene set and tissue/cell type enrichment, calculated by Data-driven Expression Prioritized Integration for Complex Traits (DEPICT) framework^31^. We prioritized the most likely ‘causal gene’ for each association using a consensus-based approach, selecting the gene with the highest, unweighted sum of evidence across all eight predictors. In the case of equality of the scores for two genes, we prioritized both genes.

### Functional annotation of genetic variants

We inferred the possible molecular consequences of genetic variants in glyQTLs. We focused on variants in LD with lead (for univariate and multivariate signals) and sentinel variants (for univariate signals) picked by COJO. We created a “long list” of putative causal variants using PLINK version 1.9 (--show-tags option), applied to whole genome re-sequenced data for 503 European ancestry individuals (1000 Genomes phase 3 version 5 data). The size of the window to find the LD in both cases was equal to 500 kb. The default value of r^2^ > 0.8 was taken as a threshold to include SNPs into the credible sets. Ensembl Variant Effect Predictor (VEP) (Supplementary Table 6e) and by FATHMM-XF (Supplementary Table 6c), FATHMM-INDEL (Supplementary Table 6d) to reveal pathogenic point mutations.

### Genes of N-glycan biosynthesis and Congenital Disorders of Glycosylation

We searched for the genes encoding glycosyltransferases – enzymes, with a known role in N-glycan biosynthesis^82^, located in the ±250 Kb-vicinity of the lead SNPs in glyQTLs. Additionally, we prioritized genes with known mutations, that cause Congenital Disorder of Glycosylation according to MedGen database (https://www.ncbi.nlm.nih.gov/medgen/76469) that are located in the vicinity of ±250 kb from the lead SNPs.

### Colocalization with eQTL and pQTL

To find potential pleiotropic effects of glyQTL on gene expression levels in relevant tissues, we applied Summary data-based Mendelian Randomization (SMR) analysis followed by the Heterogeneity in Dependent Instruments (HEIDI)^32^ on expression of quantitative trait loci (eQTLs) obtained from Westra Blood eQTL collection^83^ (peripheral blood), GTEx (version 7) eQTL collection^84^ (liver, whole blood), CEDAR eQTL collection^53^ (CD19+ B lymphocytes, CD8+ T lymphocytes, CD4+ T lymphocytes, CD14+ monocytes, CD15+ granulocytes) and on protein quantitative trait loci (pQTLs) using SomaLogic datasets^85,86^. As outcome variable we used univariate association results for the N-glycome trait with the most significant association; in the case of glyQTLs replicated only in multivariate analysis, we used summary statistics for the most associated univariate trait as the primary trait in the analysis.

The results of the SMR test were considered statistically significant if P_adj_ < 0.05 (Benjamini-Hochberg adjusted *P*). The significance threshold for HEIDI tests was set at P = 0.05 (P < 0.05 corresponds to the rejection of the pleiotropy hypothesis) (Supplementary Table 6f, 6g).

## DEPICT

Gene prioritization and gene set and tissue/cell type enrichment analyses were performed using the Data-driven Expression Prioritized Integration for Complex Traits framework (DEPICT)^31^. DEPICT analysis was conducted for SNPs associated with any N-glycosylation trait at P < 5 x 10^-8^/28 in univariate analysis and with any N-glycosylation trait group at P < 5 x 10^-8^/21 in multivariate analysis. The significance threshold for DEPICT analysis was set at False Discovery Rate FDR < 0.20 (Supplementary Table 6h, 6i, 6j).

### Colocalization with TF and IgG

In this study, colocalization analysis (SMR-θ)^53^ was conducted for total plasma, IgG and TF glyQTLs. The analysis was restricted to loci that were a) previously implicated in TF GWAS^42^ (4 loci); IgG GWAS^40^ (15 loci); both (2 loci) (Supplementary Table 5d), b) reached genome-wide significance in the GWAMA of European descent (N=10,172), c) replicated in this study. Statistic θ is a weighted correlation, whose computation requires information on p-values and effect direction. The high absolute value (e. g. |0| > 0.7) means the locus likely has a pleiotropic effect on investigated traits.

### Pleiotropy with disease

To study potential pleiotropic effects on a range of traits associated with various medical conditions SMR/HEIDI analysis was carried out similarly to that for colocalization with eQTL and pQTL.

Summary statistics for complex and medical conditions-related traits were obtained from the UK Biobank^87^, the CARDIoGRAM Consortium (http://www.cardiogramplusc4d.org/), the Psychiatric Genomics consortium (https://pgc.unc.edu/) and other trait collections from other studies (see Supplementary Table 9 for the full list of the traits analyzed). We conducted analysis separately for the disease-related traits and other complex traits.

### Associations between PGS for plasma N-glycosylation traits and disease phenotypes

To test the associations between the 117 human plasma N-glycosylation traits and ICD-10 disease phenotypes, we used logistic regression considering PGS for each glycan trait as a predictor for each disease phenotype in turn.

The list of the diseases was taken from medical histories and questionaries obtained from non-related UK Biobank participants of European descent for which we had PGS for N-glycosylation traits calculated (N = 374,303). All medical codes were preliminary filtered by prevalence (> 0.5% and < 99.5%). For this analysis we used 167 groups of codes that fall into Chapters I-XV of the UK Biobank classification of phenotypes. These codes describe a wide range of phenotypes including infectious diseases, endocrine, nutritional and metabolic diseases, diseases of the nervous system, diseases of the circulatory, respiratory, digestive and other systems, etc.

To perform logistic regression analyses we used the standard glm() function in R v.4.2.2. programming language. We included sex, age, batch number and first ten principal components of the kinship matrix (PC 1-10) as covariates in addition to the PGS predictor. Finally, we filtered out the results not passing the significance threshold for the association of P < 0.05/(28 x 167) = 1.07 x 10^-5^, where 28 is the number of plasma N-glycome principal components explaining over 99% of the 117 N-glycosylation traits variation, and 167 is the numbers of ICD-10 codes.

### Mendelian Randomization and Sensitivity Analyses

In the previous step we identified 64 pairs of associated disease phenotypes and plasma N-glycosylation traits. To investigate the causal relationships between these traits we performed a bidirectional two-sample MR analysis^33^: for each pair we performed two MR analyses using the glycosylation trait as exposure and the disease as outcome and *vice versa*.

As the sources of the summary statistics for MR analyses, we used the largest available GWAMA for plasma N-glycosylation traits in a cohort of European descent described previously in the current study (N = 10,172) and GWAS available from the UK Biobank database (for more details about these cohorts see Supplementary Table 14).

The framework of the two-sample MR was specified before the analysis. Genetic IVs for the two sample MR were identified as follows. First, the set of SNPs present both in the GWAS for the exposure and outcome traits was selected. Then for this overlapping set of SNPs in the GWAS for the exposure trait we performed clumping for independence using PLINK2^81^ within a 10,000 kb window. Additional parameters for clumping included an r^2^ > 0.001 threshold for correlation, IVs with minor allele frequency MAF < 0.05 were excluded. When plasma N-glycosylation traits were considered as exposures, *P* threshold for clumping was defined as 5 x 10^-8^/28 = 1.79 x 10^-9^ (28 - number of plasma N-glycome principal components explaining over 99% of the 117 N-glycosylation traits variation). When the disease phenotypes were considered as exposure, this threshold was set at 5 x 10^-8^/14 = 3.57 x 10^-9^ (14 - number of disease phenotypes significantly associated with at least one plasma N-glycosylation trait in the logistic regression analysis).

Summary statistics for IVs in the exposure and outcome GWAS data were processed using the TwoSampleMR R package^33^: the data were harmonized excluding ambiguous/triallelic SNPs. Only the pairs where at least 2 IVs were available for the exposure trait were considered for the further analysis. MR analysis was performed using mr_report() function from the TwoSampleMR R package. Significance thresholds for the MR results were set as 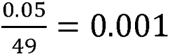 for the analysis of 49 traits pairs where glycans were considered as exposures, and as 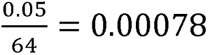 for the analysis of the 64 pairs where diseases were considered as exposures. If at least one of the MR methods used (Inverse variance weighted, MR Egger, Weighted median, Weighted mode, or Simple mode) produced a statistically significant causal estimate, that pair of traits was selected for the follow-up sensitivity analyses.

Follow-up sensitivity analyses included those automatically implemented in the mr_report() function, such as heterogeneity tests, test for directional horizontal pleiotropy, leave-one-out analysis, forest plot and funnel plot.

In addition to the sensitivity analyses described above, for the pairs where plasma N-glycosylation traits were used as exposures, since the number of IVs was very low (2-3 SNPs), we performed colocalization analysis (SMR-HEIDI) for each of the IVs. The results of the SMR test were considered statistically significant if Benjamini-Hochberg adjusted P < 0.05. The significance threshold for HEIDI tests was set at P = 0.05 which corresponds to the rejection of the pleiotropy hypothesis.

For the pairs where the disease was the upstream exposure trait (disorders of lipoprotein metabolism and other lipidaemias, E78) and 27 IVs were available, we identified pleiotropic IVs using MR-PRESSO R package, detecting the IVs for which *P* of the test for outliers in MR-PRESSO were < 1. Then we repeated the MR analysis as described above excluding these SNPs.

## Data availability

The full genome-wide summary association statistics for the 117 N-glycome traits will be made publicly available **upon publication of the paper**. The data generated in the secondary analyses of this study are included with this article in the Supplementary Tables.

## Data Availability

The full genome-wide summary association statistics for the 117 N-glycome traits will be made publicly available upon publication of the paper.

## Acknowledgements

The work of S.Sh., A.T., D.M., A.S., Y.S.A. was supported by the Research Program at the Moscow State University (MSU) Institute for Artificial Intelligence. The study was conducted using the UK Biobank resource under application #59345. The work of E.E., Y.A.T was supported by the budget project of the Institute of Cytology and Genetics FWNR-2022-0020. European Community’s Seventh Framework Programme funded project PainOmics (602736). TwinsUK is funded by the Wellcome Trust, Medical Research Council, Versus Arthritis, European Union Horizon 2020, Chronic Disease Research Foundation (CDRF), Zoe Ltd and the National Institute for Health Research (NIHR) Clinical Research Network (CRN) and Biomedical Research Centre based at Guy’s and St Thomas’ NHS Foundation Trust in partnership with King’s College London. The TwinsUK Study was approved by London-Westminster Research Ethics Committee (REC reference EC04/015), and Guy’s and St Thomas’ NHS Foundation Trust Research and Development (R&D). The TwinsUK BioBank was approved by the HRA - Liverpool East Research Ethics Committee (REC reference 19/NW/0187), IRAS ID 258513. All participants provide written, informed consent. We thank Toma Keser, Mirna Šimurina, Marija Vilaj, Jerko Štambuk, Ivan Gudelj, Thomas S. Klarić, Jasminka Krištić, Jelena Šimunović, Julija Jurić, Ana Momčilović, Najda Rudman, and Maja Hanić for their assistance with glycan analysis.

## Author contributions

S.Sh. coordinated this study; S.Sh., A.T., O.Z., D.M., A.S., E.E., E.T., A.N., S.F., N.A.P., Y.A.T. contributed to the design of the study, carried out statistical analysis; A.T., D.M., A.S., E.T., O.Z., S.Sh. produced the figures; S.Sh., A.T., O.Z., D.M., A.S., Y.A.T., contributed to interpretation of the results; S.Sh., A.T., O.Z., D.M., A.S., A.N. and Y.S.A. wrote the first version of the manuscript; S.Sh., A.T., O.Z., D.M., A.S., and Y.S.A. wrote the revised second version of the manuscript; S.Sh., E.T., E.E., S.F., V.V. and Y.A.T. contributed to data harmonization and quality control; F.V., I.T.-A., T.Š. contributed to plasma N-glycome measurements and quality contol; M.M., T.S. analyzed TwinsUK dataset and contributed to interpretation of the results; M.T., M.D. analyzed SOCCS dataset and contributed to interpretation of the results; L.K., F.W., D.P., J. Van Z., M.A. designed PainOmics study and contributed to interpretation of the results; K.S. analyzed QMDiab dataset and contributed to interpretation of the results; M.G. designed CEDAR study and contributed to interpretation of the results; C.W. and M.B.S. contributed to the data collection and analyses of EPIC-Potsdam and to the interpretation of results; Y.S.A. and G.L. conceived and oversaw the study, contributed to the design and interpretation of the results; all co-authors contributed to the final manuscript revision.

## Competing interests statement

Y.S.A. is a full-time employee of GSK PLC and receives salary and stock options as compensation. G.L. is a founder and owner of Genos Ltd, a biotech company that specializes in glycan analysis and has several patents in the field. O.Z., T.Š., F.V. and I.T.-A are employees of Genos Ltd. All other authors declare no conflicts of interest. Other authors declare no competing financial interests.

## Notes

### Author Declarations

TwinsUK: The TwinsUK Study was approved by London-Westminster Research Ethics Committee (REC reference EC04/015), and Guy's and St Thomas' NHS Foundation Trust Research and Development (R&D). The TwinsUK BioBank was approved by the HRA - Liverpool East Research Ethics Committee (REC reference 19/NW/0187), IRAS ID 258513. All participants provide written, informed consent. EPIC-Potsdam: All participants gave written informed consent for biomedical research, and the study was approved by the Ethics Committee of the State of Brandenburg, Germany. PainOmics: The study was firstly approved by the Institutional Review Boards of IRCCS Foundation San Matteo Hospital Pavia and then by the Institutional Review boards of all clinical centers (King's College London, ZOL Genk/Lanaken, St. Catherine Specialty Hospital) that enrolled patients. Copies of approvals were provided to the European Commission before starting the study. Written informed consent was obtained from all participants. SOCCS: All participants gave written informed consent and study approval was from the MultiCentre Research Ethics Committee for Scotland and Local Research Ethics committee. SABRE: All participants gave written informed consent. Approval for the baseline study was obtained from Ealing, Hounslow, and Spelthorne, Parkside and University College London research ethnics committees. QMDiab: The initial study was approved by the Institutional Review Boards of HMC and Weill Cornell Medicine-Qatar (WCM-Q) (research protocol #11131/11). Written informed consent was obtained from all participants. CEDAR: The experimental protocol was approved by the ethics committee of the University of Liege Academic Hospital.

### Summary of Updates

Author affiliations and ORCID updated.

